# Better immediate declarative memory is associated with forgetting during locomotor adaptation in chronic stroke and in older adults

**DOI:** 10.64898/2026.06.16.26355404

**Authors:** S Lipior, Y Yu, ML Kelly, AR Cain, N Schweighofer, KA Leech

## Abstract

Sensorimotor adaptation is a motor learning process that contributes to movement flexibility and is thought to arise from the interaction of fast and slow adaptive processes. Evidence suggests that declarative memory contributes to adaptation through its influence on the fast process. Although adaptation deficits are common following stroke, the mechanisms underlying these deficits remain unclear. This study investigated differences in locomotor adaptation rate and forgetting between individuals with chronic stroke and age-matched controls and examined how these measures were associated with immediate declarative memory performance. Individuals with chronic stroke (n = 23) and age- and education-matched controls (n = 21) completed four 4-minute bouts of split-belt treadmill adaptation separated by rest breaks. Adaptation rate, adaptation magnitude, and forgetting were quantified from exponential fits to normalized step-length asymmetry data. Immediate declarative memory was quantified using the Repeatable Battery for the Assessment of Neuropsychological Status, and associations between adaptation measures and immediate declarative memory were evaluated using robust linear regression. Participants with stroke adapted less (p = 0.001) and more slowly (p = 0.039) than controls during early adaptation and forgot less of the adapted behavior during the first rest break (p = 0.024). Notably, poorer immediate declarative memory performance was associated with reduced forgetting during the initial rest break, irrespective of group assignment (p = 0.035). This relationship supports the hypothesis that declarative memory contributes to adaptation through a cognitively mediated fast process. These findings suggest that cognitive impairment contributes to altered adaptation following stroke and highlight the importance of considering cognitive factors when investigating motor learning mechanisms and rehabilitation outcomes in neurological populations.

## Introduction

Motor learning, defined as a sustained change in motor behavior,^1^ is an essential component of neurorehabilitation to improve function following a stroke. An important neural mechanism driving this process is sensorimotor adaptation, a behavioral phenomenon during which motor commands are systematically updated to reduce performance errors introduced by altered environmental changes (i.e., perturbations) or bodily demands.^2,3^ This error reduction process enables flexible modification of movement over relatively short timescales (e.g., seconds to minutes versus days to weeks). Although individuals post-stroke remain capable of adapting their movements in response to perturbations,^4,5^ adaptation rate is reduced relative to controls.^6–8^ Because successful rehabilitation often requires individuals to modify existing movement patterns and acquire new ones, characterizing the processes underlying sensorimotor adaptation may provide insight into the broader mechanisms that support functional recovery following stroke.^9^

A growing body of work suggests that sensorimotor adaptation is not a unitary mechanism but rather reflects the combined contribution of multiple adaptive processes operating over distinct timescales.^10,11^ The dual-state model of sensorimotor adaptation proposes that adaptation consists of the combined contributions of a fast and slow adaptive process that operate in parallel.^8–10^ The fast process learns rapidly in response to movement errors but is forgotten quickly, whereas the slow process learns more gradually and is retained over longer timescales. As sensorimotor adaptation progresses and sensory prediction errors diminish, the contribution of the fast process decreases while the slow process continues to accumulate learning, producing the characteristic learning curve commonly observed during sensorimotor adaptation experiments, with an initially rapid change in performance followed by a slower change over time.^4,11^

Because the fast and slow processes differ in learning rate and retention,^11^ evaluations of adaptation rate and the magnitude of forgetting may provide insight into their relative contributions to the observed motor output during adaptation. Here, forgetting is defined as the loss of adapted behavior observed between the end of one adaptation bout and the beginning of the next, following a rest interval. The fast process decays exponentially within minutes when feedback is removed or passive rest delays are introduced, demonstrating its fundamental temporal volatility relative to the stable memory of the slow process.^11,13–15^ Similarly, because the fast process contributes strongly to the rapid reduction of movement errors early in adaptation, faster adaptation rates may, in part, reflect a greater contribution of the fast process. Consequently, adaptation paradigms incorporating repeated adaptation bouts separated by rest periods provide a useful experimental approach for dissociating the contributions of fast and slow learning processes.^16,17^ Understanding how these adaptive processes contribute to behavior may therefore provide insight into the mechanisms underlying adaptation in both healthy and neurologically impaired populations.

The fast process is generally considered explicit, cognitively driven, whereas the slow process appears to depend more heavily on implicit, sensory-prediction-error-driven mechanisms.^18–20^ Support for a cognitive contribution to the fast process primarily comes from studies that demonstrate the development of explicit movement strategies to reduce errors early in reaching adaptation.^19–21^ This aligns with evidence of cognitive-motor interference during early adaptation^17,18^ and reduced adaptation rate or magnitude in individuals with cognitive impairments.^22–24^ Through this work, several cognitive processes have been shown to contribute to adaptation; however, accumulating evidence suggests that declarative memory may play a particularly important role in the fast process.^25–27^ For example, individuals with mild cognitive impairment, *a condition characterized primarily by memory deficits*, exhibited selective impairments in the fast process during an upper-extremity adaptation task compared with age-and education-matched controls.^24^ Collectively, these findings suggest that memory-related cognitive processes contribute to the fast process and may therefore influence both the adaptation rate and forgetting during sensorimotor adaptation.

Despite growing evidence that cognitive processes contribute to sensorimotor adaptation, the potential contribution of post-stroke cognitive impairment to altered adaptation behavior remains poorly understood. Although cognitive deficits, including memory impairment, are common following stroke,^28,29^ slower adaptation following stroke is often attributed to motor impairments. Consistent with the possibility that altered adaptation following stroke may reflect an influence of cognitive processes, a recent study by Wood et al. reported impairment of the fast process during a locomotor adaptation paradigm following a stroke relative to controls, while the slow process was preserved.^30^ However, cognitive function was not assessed in that study. An examination of adaptation rate, forgetting, and their relationship to immediate memory abilities is needed to provide insight into the mechanisms underlying adaptation behavior following stroke.

To examine these relationships in a behavior highly relevant to functional mobility after stroke, we investigated adaptation during split-belt treadmill walking.^31,32^ This paradigm provides a unique context for evaluating cognitive contributions to adaptation because walking is typically considered less cognitively demanding than upper-extremity adaptation tasks. Consequently, evidence linking memory function to locomotor adaptation would suggest that cognitive processes contribute to adaptation even in relatively automatic motor behaviors. Therefore, the purpose of this study was to quantify adaptation rate and forgetting during split-belt locomotor adaptation in individuals with chronic stroke and age-matched controls and to determine how these adaptation behaviors relate to immediate declarative memory.

Using a split-belt treadmill paradigm with repeated adaptation blocks separated by rest periods (Figure 1A, B), we quantified locomotor adaptation rates and short-term forgetting across multiple perturbation exposures. Based on the dual-state framework and evidence of selective impairment of the fast process after stroke, we hypothesized that individuals with chronic stroke would exhibit slower initial adaptation rates but reduced forgetting across rest intervals relative to age-matched controls (Figure 1C). This prediction follows from the premise that rapid error reduction during early adaptation depends on a declarative-memory-supported fast process. As the fast process is also thought to underlie the rapid decay of adapted behavior during brief rest periods, reduced engagement of this process should manifest as diminished forgetting. Consistent with this mechanism, we further hypothesized that immediate declarative memory abilities would be positively associated with the magnitude of forgetting across all participants. Demonstrating such a relationship would provide evidence that immediate declarative memory contributes to locomotor adaptation, extending cognitive accounts of sensorimotor adaptation to a behavior traditionally viewed as largely automatic.

**Figure 1.**
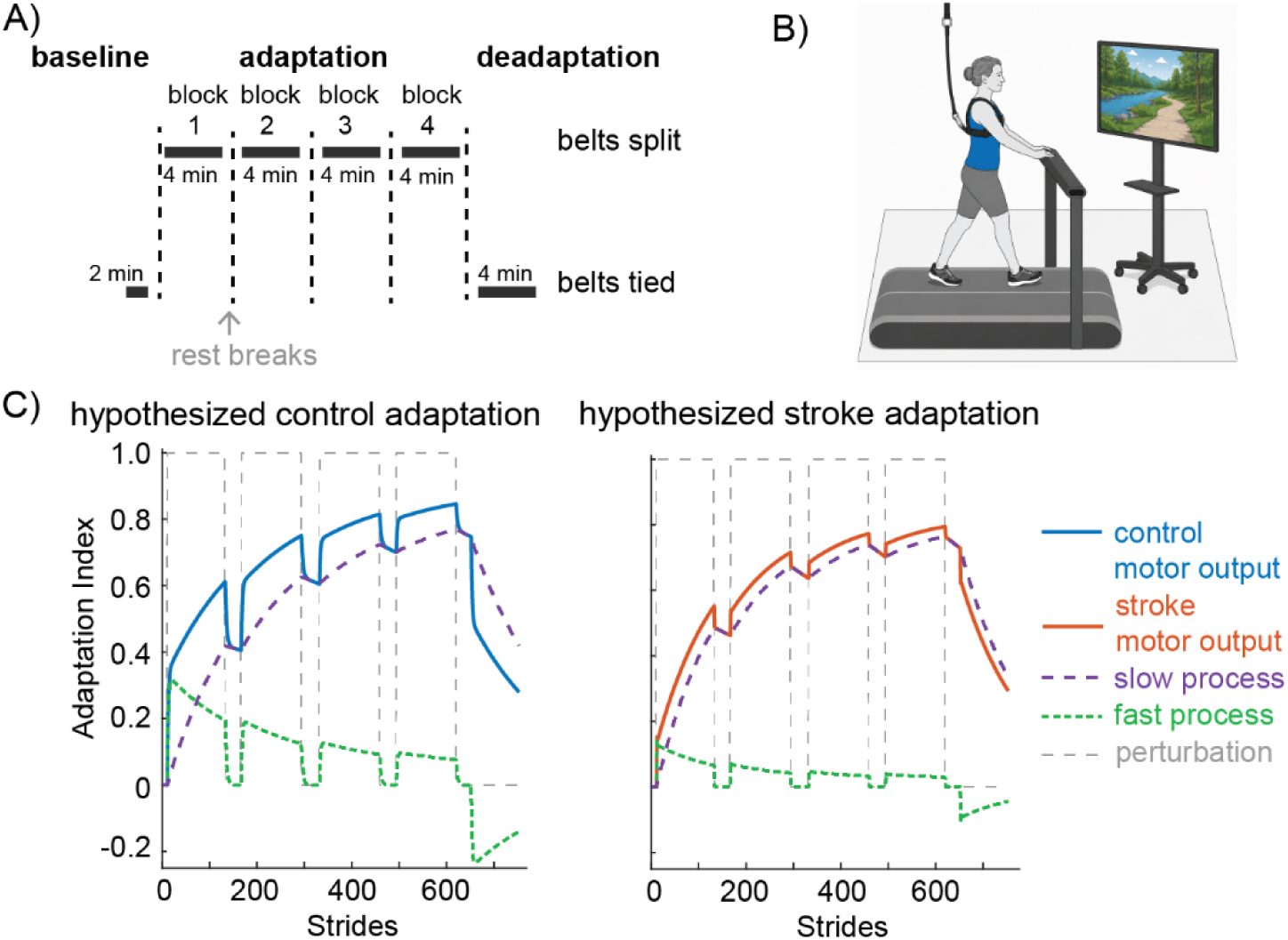
Experimental paradigm, setup, and hypothesized dual-state adaptation outcomes. A) Experimental protocol timeline illustrating the baseline phase, adaptation phase divided into four 4-minute blocks of split-belt walking separated by brief rest breaks (vertical dashed lines), and de-adaptation phase. B) Schematic of the experimental setup with a participant walking on the split-belt treadmill in a safety harness. Of note, the screen was placed directly in front of the participant during the experiment, but is displayed off to the side in the schematic to allow for visualization of the screen. C) Schematic dual-state model simulations of locomotor adaptation for neurotypical controls and individuals post-stroke. Hypothesized behavioral trajectories for neurotypical controls (left) and individuals post-stroke (right), formalized using a parallel dual-state architecture.^11,12^ The temporally volatile fast process (green dashed line) drives early error reduction but rapidly decays during rest breaks, whereas the slow process (purple dashed line) accumulates gradually and remains stable. Total motor output (solid blue/orange lines) reflects the combined contribution of both processes in response to the perturbation (grey dashed line). The stroke model illustrates a selective deficit in the fast process, manifesting as a slower initial adaptation rate and reduced step-like decay (forgetting) across rest intervals, while achieving a similar overall adaptation magnitude by the final learning block. To isolate this mechanism, the fast learning rate (B_f_ =0.14), slow learning rate (B_s_ = 0.007), and slow retention factor (A_s_ = 0.999) were held constant across models, whereas the fast-process retention factor was lower in the stroke model (A_f_ = 0.05) than in the control model (A_f_ = 0.72), using parameter values hand-tuned to qualitatively capture the experimental timeline.

## Materials and Methods

### Participants

Individuals with stroke, as well as age- and education-matched control participants, were recruited to participate in this study. To be enrolled, participants with stroke had to meet the following criteria: (1) between 45 and 85 years old, (2) unilateral, chronic (>6 months) stroke, (3) paresis confined to one side, (4) able to walk independently for five minutes without stopping. Individuals with stroke were excluded if they met the following criteria: (1) evidence of brainstem or cerebellar stroke, (2) aphasia, (3) any major musculoskeletal or non-stroke neurological condition that interferes with the assessment of sensorimotor and cognitive function, (4) uncontrolled hypertension, (5) significant cognitive deficit or dementia (<19 on MoCA). Inclusion criteria (1) and (4) and all exclusion criteria applied to the control participants. The study was approved by the University of Southern California Institutional Review Board and is registered on clinicaltrials.gov (NCT04829071). All individuals provided written informed consent prior to participation.

### Clinical Assessments

Before all other study procedures, participants completed clinical assessments of motor and cognitive function. The lower-extremity portion of the Fugl-Meyer (FM-LE) was administered to all stroke participants to assess motor impairment, with higher scores indicating less impairment.^33,34^ All participants completed the Berg Balance Scale, a measure of static balance, with higher scores indicating better balance,^35^ and the 10-meter walk test to determine their self-selected and fastest comfortable overground walking speeds.^36^ All participants also completed the Repeatable Battery for the Assessment of Neuropsychological Status (RBANS) to measure neurocognitive functioning in five cognitive domains: Immediate Memory, Visuospatial/Constructional, Language, Attention, and Delayed Memory.^37^ The RBANS provides an age- and education-normalized index score for each cognitive domain, as well as a total composite score for global cognitive function. Possible scores range from 40 to 160, with higher scores indicating better cognitive function. Here, we quantified immediate declarative memory using the RBANS Immediate Memory Index score, which assesses it via the List Learning and Story Memory subtests.

### Experimental Design

Our experimental paradigm consisted of walking on a split-belt treadmill (Woodway USA) with two belts, each driven by its own motor, allowing independent control of each belt’s speed. Prior to beginning the experiment, participants were told that the treadmill belts may move at different speeds without warning. For safety, participants were allowed to hold the front handrail and were required to wear a safety harness. The safety harness was mounted to the ceiling to prevent falls if engaged, but it did not provide body-weight support while walking.

Participants performed three phases of treadmill walking: Baseline, Adaptation, and De-adaptation (Figure 1A). During the Baseline (2 minutes) and De-adaptation phases (4 minutes), the treadmill belts moved at the same speed (half the speed of the fast belt), referred to as the tied-belt configuration. During the Adaptation phase (four 4-minute blocks; Figure 1A), the treadmill belts moved at a 2:1 speed ratio to create a split-belt configuration. Consistent with prior research on locomotor adaptation in neurologic populations,^4,5^ the fast belt speed was set to the fastest treadmill gait speed tolerated by the participant, and the slow belt speed was set to half that speed. The fastest treadmill speed was determined using a custom staircase algorithm based on their fastest comfortable overground walking speed. To avoid large differences in treadmill speeds between the stroke and control groups, the fast belt speed was set to 1.0 m/s for the control group as in previous work.^30^ All participants were provided rest breaks between the four adaptation blocks. During each break, participants could either remain standing on the stationary treadmill or sit in a chair placed on it, allowing them to rest without stepping off. Participants were allowed to rest for up to five minutes, but we did not control the amount of rest time between participants or blocks.

Average step lengths measured during Baseline walking were used to determine which belt should be the fast belt during the Adaptation phase. Consistent with prior work, the belt under the limb with the shorter average step length was the fast belt, while the belt under the limb with the longer average step length was the slow belt.^2^ This belt-speed arrangement was selected to produce a perturbation that exaggerated the baseline step length asymmetry. To limit the possible contribution of conscious step length asymmetry corrections driven by visual feedback about task performance,^38,39^ participants were instructed not to look at their feet while walking and instead to watch a recording of a nature hike on a television screen placed directly in front of the treadmill (Figure 1B).

### Kinematic Data Collection

During the experiment, participants wore a custom 12-marker set including one marker for each iliac crest (pelvis), greater trochanter (hip), lateral epicondyle (knee), lateral malleolus (ankle), great toe, and fifth metatarsal joint. Three-dimensional kinematic data was captured with 8 motion capture cameras (Miqus M3, Qualysis, Göteborg, Sweden). Marker trajectories were sampled at 100 Hz and collected via Qualysis Track Manager software (v.2021.1 Qualysis, Göteborg, Sweden).

### Kinematic Data Processing

Custom MATLAB scripts (vR2026a, MathWorks, Natick, MA) were used to analyze motion-capture data from ankle markers. Marker position data were low-pass filtered at 6 Hz. Step length was calculated as the difference between the position of the ankle marker on the leading limb and the trailing limb at heel strike.

Step length asymmetry (SLA) for each stride was calculated by normalizing the difference between the bilateral step lengths to their sum^8,30,40,41^:

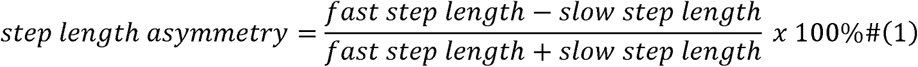

SLA values of 0 indicate perfect symmetry between the slow and fast step lengths, while larger-magnitude negative values indicated greater step length asymmetry. To normalize SLA across participants, we baseline-corrected the data by subtracting the participant’s mean SLA during the Baseline phase from each stride during the Adaptation and De-adaptation phases. Consistent with prior work,^30^ we removed the first stride of each Adaptation block to account for treadmill acceleration.

To compare adaptation behaviors across participants, we transformed the step length asymmetry data into a normalized Adaptation Index. We modified the standard calculation used in prior locomotor adaptation paradigm^30,39^ to explicitly account for instances in which participants adapt beyond their baseline asymmetry. For each participant, the Adaptation Index for each stride s was calculated as follows:

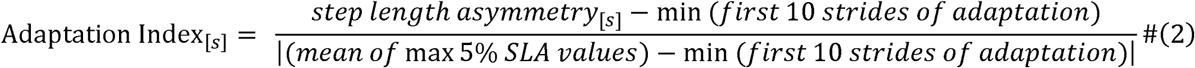

During the Adaptation phase, an Adaptation Index of 0 represents the minimum (most negative) step length asymmetry in the first 10 strides of Adaptation Block 1 and 1 represents the mean of the maximum (most positive) 5% SLA values. Therefore, 0 represents maximal perturbation for a given participant, while 1 represents complete adaptation. The equation used to scale De-adaptation data from Figure 2 is included in Supplementary Material. These data will be evaluated in a future study.

**Figure 2:**
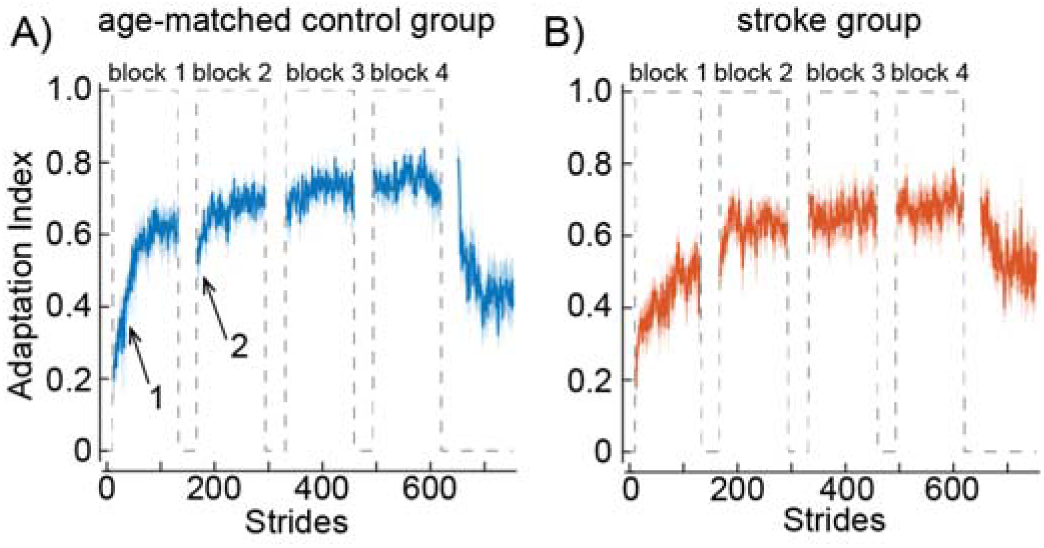
Mean adaptation index for the control group (A) and stroke group (B) throughout the experiment. Each block was truncated to the participant with the fewest strides for group-mean visualization purposes. Shading represents standard error of the mean. Arrow 1 indicates faster adaptation while arrow 2 indicates greater forgetting in the control group.

### Quantification of Adaptation Behaviors: adaptation rate, adaptation magnitude, forgetting

To robustly quantify the dynamics of locomotor adaptation and short-term forgetting, we applied single-exponential curve fits to this normalized Adaptation Index during the first two blocks. This mathematical framework provides several distinct advantages over traditional epoch-averaging techniques (e.g., averaging the first or last 5 to 10 strides). First, split-belt locomotion inherently exhibits high trial-by-trial motor variability, particularly in neurologically impaired populations; exponential fits effectively smooth this high-frequency noise by leveraging the entire behavioral curve rather than relying on isolated, high-variance data points.^11,42^ Second, continuous exponential modeling eliminates the arbitrary bias introduced by selecting fixed trial windows, which can artificially distort estimates if a participant adapts rapidly within the first few strides.^43^

Adaptation behavior within each block was characterized using a single-exponential model of the form:

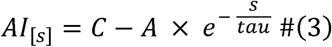

where *AI_[s]_* was the Adaptation Index for stride *s*, *C* is the asymptotic value, *A* is the amplitude, and *tau* is the time constant indicative of the adaptation rate. Smaller values of *tau* indicate faster adaptation toward the asymptote.

From these exponential fits, we extracted three distinct behavioral measures: early adaptation rate, early adaptation magnitude, and forgetting magnitude. Early adaptation rate was quantified using the natural logarithm of the estimated time constant (*tau*) from Block 1; because exponential rate parameters typically exhibit a right-skewed distribution, this log-transformation normalizes the data for parametric statistical testing. Early adaptation magnitude was calculated as the difference between the model-estimated adaptation levels at the end versus the beginning of Block 1. Specifically, adaptation magnitude was calculated as:

Adaptation magnitude = AI_Bl,end_ - AI_Bl,start_ , where AI_Bl,start_ represents the initial value of the fitted exponential curve for Block 1 and AI_Bl,end_ represents the final value of the fitted exponential curve for Block 1. The same calculation was also performed for Block 2. Finally, forgetting was quantified as the discrete difference between the final value of the fitted curve for Block 1 and the initial value of the fitted curve for Block 2. Specifically, forgetting was calculated as: Forgetting = AI_Bl,end_ - AI_B2,start_ , where AI_Bl,end_ represents the final value of the fitted exponential curve for Block 1 and AI_B2,start_ represents the initial value of the fitted exponential curve for Block 2. On this scale, more positive values indicate a greater loss of adapted behavior across the rest interval (i.e., greater forgetting). By using model-estimated projections rather than raw stride averages as in previous locomotor adaptation studies,^44–46^ this calculation isolates pure motor decay from the immediate step-to-step noise of the rest-to-walk transition, ensuring a robust behavioral representation of the temporally volatile, fast process.^47^

The model was fitted separately to each individual participant’s Adaptation Index data from Blocks 1 and 2 using MATLAB’s *fmincon* global optimization function. Model parameters were estimated by minimizing the sum of absolute residuals between the observed and predicted Adaptation Index values. The *C* and *A* parameters were constrained to be between 0 and 1, while the *tau* parameter was constrained to be between 5 and 550 to prevent implausibly small or large values of learning rate.

### Statistical Analysis

Statistical analyses comparing demographics between the groups were conducted in RStudio (v4.5.1). All other statistical analyses were performed in MATLAB (vR2026a). To compare the demographic characteristics of the stroke and control groups, we performed independent-samples t-tests. Behavioral data normality was assessed using the Shapiro-Wilks’ test. For variables that satisfied assumptions of normality (forgetting), group differences were evaluated using independent samples t-tests. Normally distributed data were expressed as mean ± SD. For variables that violated normality assumptions (Shapiro-Wilks’ p<0.05; adaptation magnitude and log of *tau*), group differences were expressed as median with interquartile ranges (IQR), 25% - 75%, and compared using the Mann-Whitney test for independent measures. To determine if the relationship between memory and adaptation behaviors (forgetting and adaptation rate) was different between the stroke and control group, we ran separate regressions with interaction term for Immediate Memory x Group. Because we found that these interactions were not significant for forgetting or adaptation rate, we combined the groups for subsequent analyses. Associations between immediate memory and adaptation parameters were examined using robust linear regression. We used a robust linear regression for these analyses because we identified influential points in the data using Cook’s distance, with observations exceeding a threshold of 4/n (n = sample size) considered potentially influential. We used robust linear regression to reduce the impact of these observations. The significance level was set at α = 0.05 for all analyses.

## Results

We enrolled 24 stroke survivors and 23 control participants in this study. On average, the control participants were of a similar age (stroke = 57.3 ± 8.3 years, control = 60.1 ± 6.5 years; p = 0.22) and level of education (stroke = 15.2 ± 2.6 years, control = 16.0 ± 2.4 years; p = 0.32) as the stroke participants. Of the 24 individuals recruited for the stroke group, we excluded 1 from the analysis because they were unable to complete the split-belt walking task due to fatigue. Of the 23 individuals recruited for the control group, two were excluded because English was their second language and cognitive testing was not conducted in their primary language.

Furthermore, one control participant was excluded from the analyses of immediate memory because their immediate memory index score (16) was an outlier, falling 5.6 standard deviations below their age- and education-normative mean (standard score = 100), whereas all other participants in both groups were within 3.1 standard deviations of their normative mean. Given the magnitude of the deviation, this was considered unlikely to be representative of the study population. The demographics and clinical characteristics of participants in each group included in the analyses are shown in Supplementary Table 1. Of note, the RBANS total scores were significantly lower in the stroke group (85.1 ± 15.6) compared to the control group (101.3 ± 15.7; p = 0.001). Immediate memory index scores followed a similar pattern, with the stroke group scoring significantly lower (87.1 ± 18.7) than the control group (100.8 ± 23.6; p = 0.04).

### Adaptation behavior differed between the stroke and control groups

Figure 2 displays the mean adaptation index for the age-matched control (A) and stroke (B) groups during the four adaptation blocks and the de-adaptation block of the experiment. In both groups, introduction of the split-belt perturbation produces an immediate increase in asymmetry followed by gradual adaptation. We observed a difference in early adaptation between the groups: faster adaptation (Figure 1A; arrow 1) and more forgetting between blocks 1 and 2 (Figure 1A; arrow 2) in controls relative to the stroke group.

To quantify these differences, we analyzed the adaptation index in blocks 1 and 2. Using the exponential model, we estimated adaptation magnitude, rate, and forgetting magnitude during early adaptation. Representative model fits for individual participants from each group are shown in Figure 3A and B. These examples demonstrate the ability of the exponential model to well characterize individual adaptation behavior and quantify adaptation magnitude, adaptation rate, and forgetting magnitude across the first rest interval. Overall, the exponentials provided adequate fit to participant adaptation behavior, with a Block 1 RMSE of 0.121 ± 0.045 (mean ± SD) for controls and 0.172 ± 0.089 for post-stroke subjects. Similarly, Block 2 RMSE was 0.102 ± 0.034 for controls and 0.152 ± 0.082 for post-stroke subjects. All individual model fits are provided in Supplemental Figures 1 and 2.

**Figure 3:**
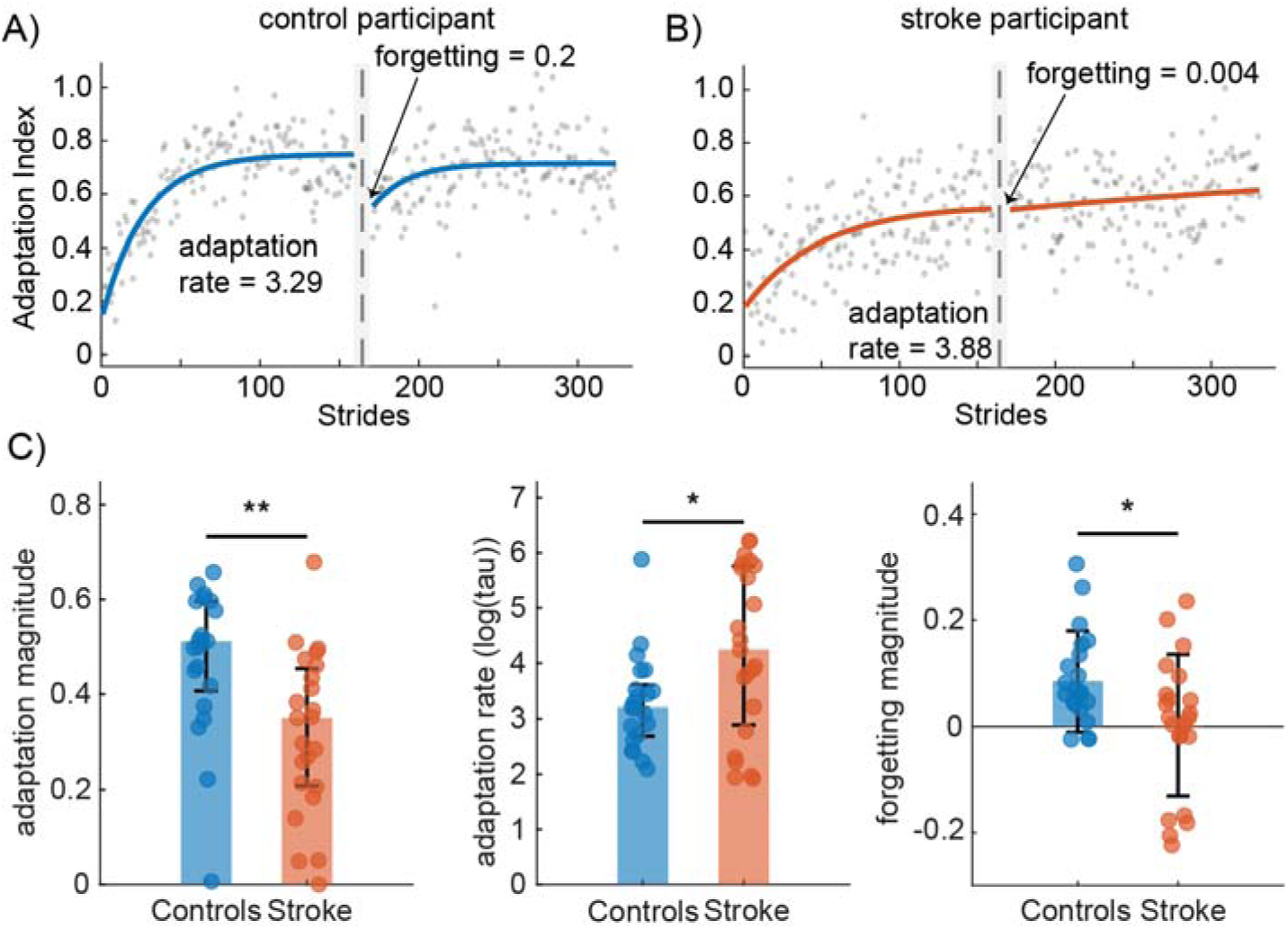
Individual exponential model fits of adaptation behavior and group adaptation metrics. Representative individual exponential model fits from a post-stroke participant (A) and a control participant (B). The representative control participant shows faster adaptation rate (3.29) and more forgetting (0.2) while the representative stroke participant shows slower adaptation rate (3.88) and less forgetting (0.004). (C) Group comparisons of adaptation magnitude (median±IQR), adaptation rate (median±IQR), and forgetting (mean±SD) values derived from individual exponential model fits. On average, individuals post-stroke demonstrated reduced adaptation magnitude, slower adaptation, and less forgetting across the first rest interval relative to control participants. Individual participant data shown as overlaid data points. * p < 0.05, ** p < 0.01.

Adaptation behavior differed between the post-stroke and control groups (Figure 3C). Adaptation magnitude during Block 1 was smaller in the stroke group (0.351, IQR = [0.208-0.454]) relative to controls (0.511, IQR = [0.407-0.596]; p = 0.001). The post-stroke group also adapted more slowly (larger log of *tau*) during Block 1 (4.241, IQR = [2.891-5.761]) than the control group (3.203, IQR = [2.689-3.606]; p = 0.039). Forgetting magnitude across the first rest interval was also lower in the post-stroke group (0.002 ± 0.133) than in the control group (0.085 ± 0.095; t(42) = -2.35; p = 0.024), indicating that the control group forgot more of the adapted behavior during the first rest interval. The groups did not differ significantly in adaptation magnitude (p=0.47) or rate (p=0.12) in Block 2.

### Immediate memory scores were positively associated with forgetting in early adaptation

To examine the relationship between memory and adaptation behaviors, we first tested whether the relationship between immediate declarative memory and forgetting magnitude differed between control and post-stroke participants. The Immediate Memory × Group interaction was not significant (*p* = 0.809), indicating a similar relationship across groups. The same analysis for adaptation rate also revealed no significant interaction (*p* = 0.905). Therefore, subsequent analyses were conducted on the combined sample.

We examined the associations between immediate memory scores and adaptation rate and forgetting magnitude in separate robust linear regression analyses. There was a significant positive correlation between immediate memory and forgetting (beta = 0.002, t = 2.175, p = 0.035; Figure 4). That is, participants with higher immediate memory scores exhibited a greater forgetting magnitude (i.e., a more positive change in the Adaptation Index) between Blocks 1 and 2 than those with poorer memory. There was no significant relationship between immediate declarative memory and adaptation rate (beta = -0.01, t = -1.091, p = 0.282).

**Figure 4:**
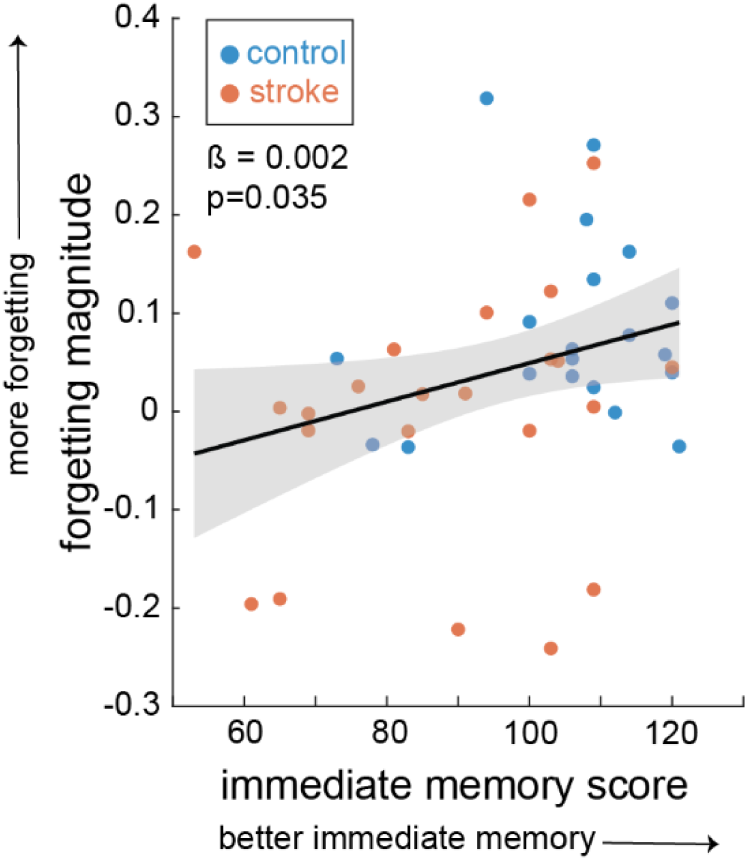
Relationship between forgetting magnitude and immediate declarative memory. Scatter plot showing the association between immediate memory index score from the RBANS and forgetting magnitude across all participants. Blue circles represent age-matched controls (*n* = 20), and orange circles represent individuals post-stroke (*n* = 23). The solid black line indicates the robust regression fit, and the shaded area represents the 95% CI of the model.

## Discussion

The purpose of this study was to investigate differences in adaptation rate and forgetting during split-belt locomotor adaptation between individuals with chronic stroke and age-matched controls and to determine whether these adaptation behaviors were related to immediate declarative memory performance. Consistent with our hypothesis, individuals with stroke demonstrated slower adaptation and reduced forgetting across rest periods relative to controls, suggesting diminished engagement of the fast process and greater reliance on a slower, more stable learning mechanism. Notably, we found an association between Immediate Memory Index scores and adaptation behavior across all participants: individuals with *better immediate declarative memory* exhibited *greater forgetting* between the first two adaptation bouts. Although this finding appears counterintuitive, it is consistent with the dual-state model of sensorimotor adaptation and provides additional evidence that memory-related cognitive processes contribute to the fast process.

Our finding that poorer immediate declarative memory was associated with reduced forgetting across rest intervals suggests that individuals with lower memory abilities may be less able to recruit this cognitively mediated, temporally labile process, resulting in a greater relative contribution of the slower, more stable process and consequently less forgetting. This interpretation is consistent with previous arm visuomotor adaptation data and computational work demonstrating that forgetting and interference disproportionately affect the fast process, thereby shifting learning toward slower, more durable mechanisms.^10^ These findings also reinforce observations from our previous work on motor skill learning after stroke.^48^ In that study, individuals with intact visuospatial working memory exhibited the classic contextual interference effect, showing less long-term forgetting after random than blocked practice. In contrast, individuals with poorer working memory were largely insensitive to the practice schedule and demonstrated stable retention under both conditions. We interpreted these findings within a fast/slow learning framework, proposing that individuals with greater cognitive capacity rely more heavily on a fast but fragile learning process, whereas individuals with reduced cognitive capacity depend more on slower, more stable learning mechanisms. The present results extend this account to locomotor adaptation by demonstrating the same relationship between immediate memory capacity and forgetting over much shorter timescales. Together, these findings suggest that memory capacity may be an important determinant of the relative contribution of fast and slow learning processes across multiple forms of motor learning.

These results also extend prior observations linking declarative memory to sensorimotor adaptation or motor skill learning beyond upper-extremity reaching tasks. Much of the evidence supporting a cognitive contribution to adaptation has emerged from reaching paradigms,^9,19^ where the development of explicit movement strategies (e.g., conscious aiming^49^) is thought to play a prominent role in early learning. Consequently, it has remained unclear whether relationships between memory function and adaptation reflect a general property of sensorimotor adaptation or are specific to the cognitive demands of reaching tasks. Locomotor adaptation provides an important context for addressing this question because walking is often considered a relatively automatic motor behavior. The observed association between immediate declarative memory performance and forgetting during locomotor adaptation supports the notion that interactions between declarative memory and sensorimotor adaptation are not restricted to upper-extremity reaching tasks and may instead represent a more general feature of adaptive motor learning.

Our results provide an interesting contrast to findings from visuomotor adaptation research that has emphasized the role of visuospatial working memory in early adaptation behavior.^50–52^ One possible explanation is that different adaptation paradigms place distinct cognitive demands on the learner. Reaching adaptation tasks often require participants to use visual information to formulate and maintain explicit aiming strategies, placing substantial demands on visuospatial working memory. In contrast, split-belt locomotor adaptation occurs during continuous walking and appears to be at least partially dissociable from visually-guided gait corrections.^39,53^ Consequently, the cognitive resources supporting locomotor adaptation may differ from those recruited during visuomotor adaptation tasks. Although speculative, these findings suggest that cognitive engagement may be a general feature of sensorimotor adaptation across motor behaviors, while the specific cognitive processes that support adaptation may vary according to the sensory and task demands of the behavior being adapted.

Our results are consistent with prior evidence from several lines of research suggesting that cognitive processes contribute to locomotor adaptation. Dual-task studies have shown different patterns of cognitive-motor interference when locomotor adaptation paradigms are conducted with simultaneous working memory tasks.^38,40,54,55^ A recent pilot study has also demonstrated that individuals with mild cognitive impairment adapt less while walking than age-matched controls.^22^ Collectively, these studies indicate that immediate memory processes contribute to locomotor adaptation, with our results highlighting the role of immediate declarative memory in early locomotor adaptation.

These observations have important implications for understanding the mechanisms underlying altered sensorimotor adaptation after stroke. While individuals with stroke demonstrated slower adaptation and reduced forgetting than controls, the relationship between immediate declarative memory performance and forgetting was observed across all participants rather than being specific to the stroke group. This pattern suggests that adaptation behavior is influenced not only by the integrity of sensorimotor systems but also by cognitive processes related to declarative memory. The inclusion of a control group allowed us to partially dissociate the effects of stroke-related motor impairment from cognitive influences on adaptation, demonstrating that memory performance explained variability in adaptation behavior across both groups. These findings support a broader framework^56^ in which adaptive motor behavior emerges from interactions between sensorimotor and cognitive learning mechanisms and may help explain individual differences in adaptation that are not accounted for by motor impairment alone.^57^

This work has several potential implications for neurorehabilitation following stroke. Sensorimotor adaptation is considered one of the more automatic motor learning processes,^1^ placing fewer demands on cognitive resources than other motor learning processes. However, our results indicate that individuals with memory impairments may have a reduced capacity for rapid adaptation to novel movement demands and may therefore require longer or repeated exposure to a perturbation to achieve similar levels of adaptation than previously appreciated. More broadly, our results suggest that cognitive function, particularly memory performance, may contribute to the substantial variability in motor learning outcomes observed after stroke.

Although additional work is needed to determine how cognitive impairments influence responsiveness to rehabilitation interventions, a better understanding of the mechanisms underlying motor learning may ultimately support the development of more personalized rehabilitation strategies that account for an individual’s cognitive-motor profile and optimize treatment delivery.

## Conclusion

In this study, individuals with chronic stroke demonstrated slower adaptation, lower adaptation magnitude, and less forgetting than age-matched controls during the early stages of split-belt locomotor adaptation. Across all participants, poorer immediate declarative memory was associated with reduced forgetting between the first two blocks of adaptation, a finding consistent with diminished engagement of the fast process, as predicted by the dual-state model of sensorimotor adaptation. These findings support a role of immediate declarative memory in locomotor adaptation and provide new insight into the mechanisms underlying altered adaptation after stroke, suggesting that cognitive impairment may contribute to adaptation deficits in this population.

## Data Availability

All data produced in the present study are available upon reasonable request to the authors

## Conflicts of Interest and Source of Funding

The authors report no conflicts of interest. This work was supported by NIH Grant K01 AG073467. This work was supported by the National Center for Advancing Translational Science (NCATS) of the National Institute of Health under award number UL1TRR001855. The content is solely the responsibility of the authors and does not necessarily represent the official views of the National Institutes of Health.

This study is registered on clinicaltrials.gov (NCT04829071).

