## Supplementary Materials for "Better immediate declarative memory is associated with forgetting during locomotor adaptation in chronic stroke and in older adults"

**Table 1. Demographics and clinical characteristics of participants in the stroke and control groups.**

|  | Stroke  (n=23) | Control  (n=21) | p value |
| --- | --- | --- | --- |
| Age (years) | 57.3 ± 8.3 | 60.1 ± 6.5 | 0.22 |
| Biological Sex | 8F/15M | 7F/14M | -- |
| Time since stroke (months) | 83.6 ± 71.2 | -- | -- |
| More affected side | 13L/10R | -- | -- |
| Lower extremity Fugl-Meyer | 23.4 ± 4.8 | -- | -- |
| Years of education | 15.2 ± 2.6 | 16.0 ± 2.4 | 0.32 |
| RBANS total score | 85.1 ± 15.6 | 101.3 ± 15.7 | 0.001** |
| RBANS immediate memory | 87.1 ± 18.7 | 100.8 ± 23.6 | 0.04* |
| Self-selected overground walking speed (m/s) | 0.75 ± 0.26 | 1.2 ± 0.21 | <0.001*** |
| Fastest comfortable overground walking speed (m/s) | 1.1 ± 0.4 | 1.8 ± 0.31 | <0.001*** |
| Fast treadmill belt speed (m/s) | 0.98 ± 0.33 | 1.0 ± 0 | 0.72 |

All continuous variables are reported as the means ± standard deviations. P-values reflect results from independent-samples t-tests comparing the groups. * indicates p < 0.05, ** indicates p < 0.01, *** indicates p < 0.001. Abbreviations: F, female; M, male; R, right; L, left; m/s, meter/second; RBANS, Repeatable Battery for the Assessment of Neuropsychological Status.

**De-adaptation equation**:

$$\begin{aligned} {Adaptation Index}_{\left[ s \right]}= \frac{{step length asymmetry}_{[s]}-(mean of\min5\% SLA values)}{\left| (mean of\min5\% SLA values)-\max(first 10 strides of deadaptation) \right|} \# \end{aligned}$$

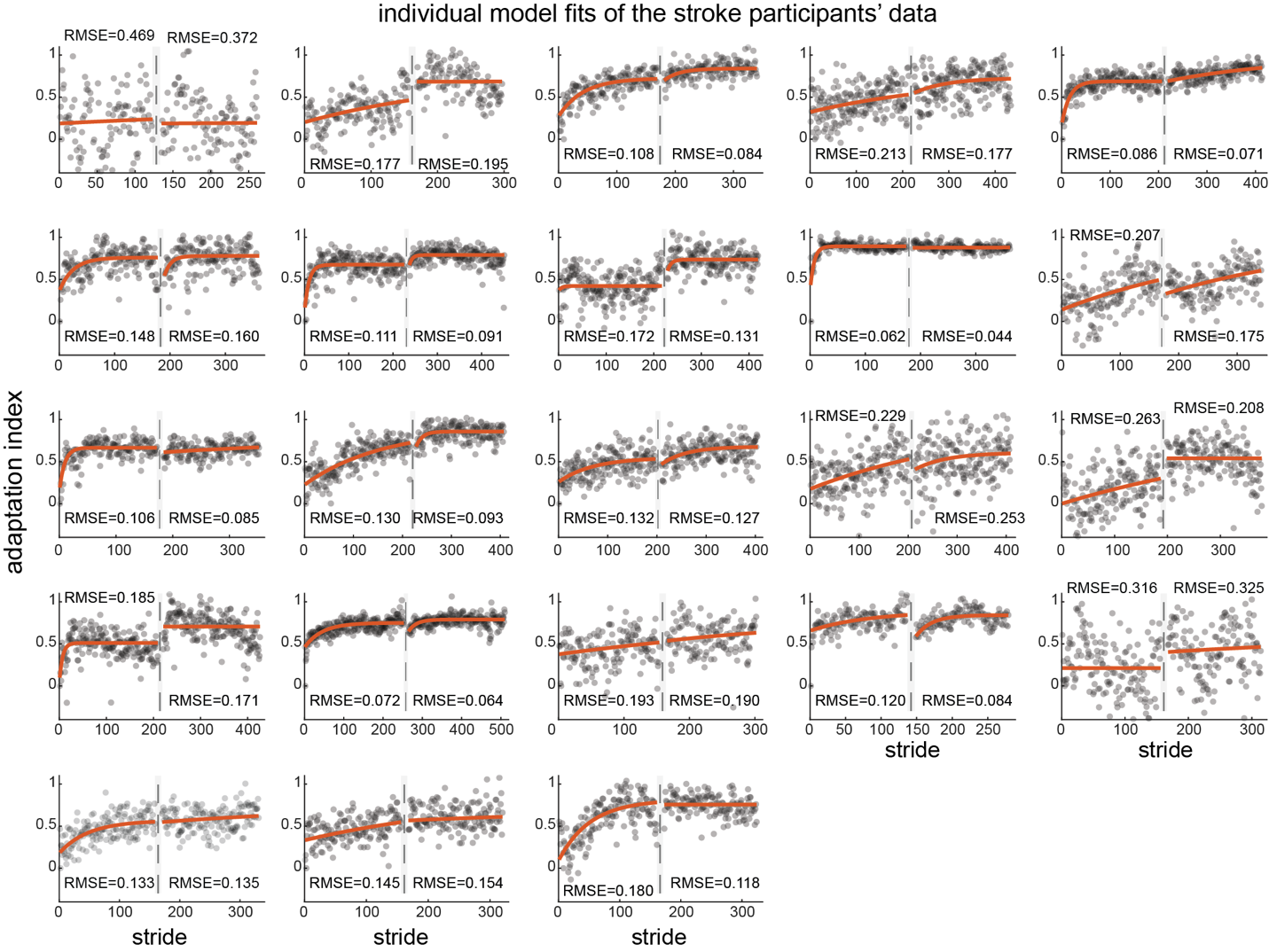

**Supplementary Figure 1: Individual exponential model fits for post-stroke participants.** Each panel displays the Adaptation Index values from a single participant. Gray circles represent stride-by-stride Adaptation Index values during Adaptation Blocks 1 and 2, separated by a dashed vertical line indicating the rest interval. Orange lines represent the fitted exponential models. RMSE values for Block 1 and 2 are displayed adjacent to the corresponding model fit.

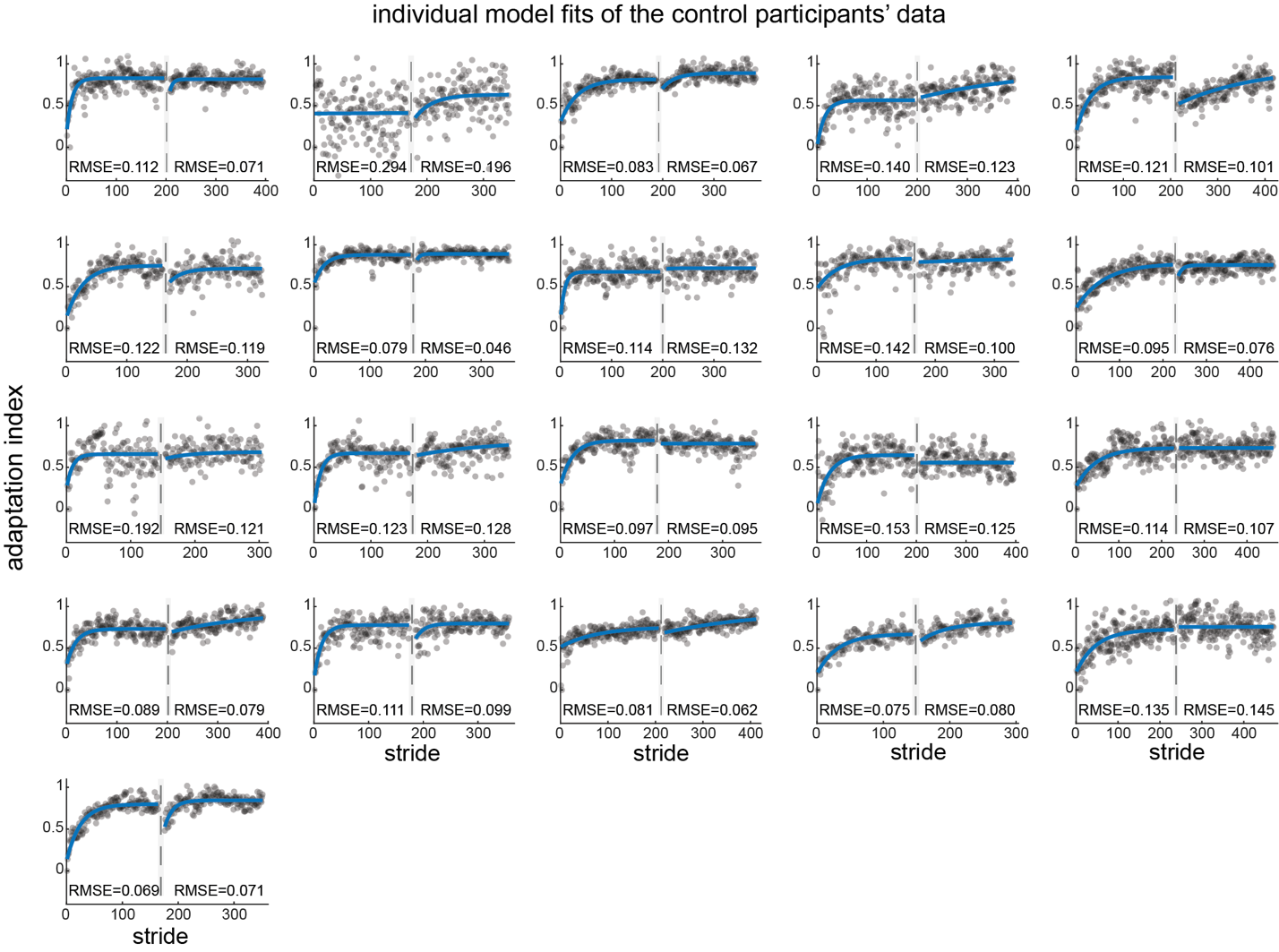

**Supplementary Figure 2: Individual exponential model fits for control participants.** Each panel displays the Adaptation Index values from a single participant. Gray circles represent stride-by-stride Adaptation Index values during Adaptation Blocks 1 and 2, separated by a dashed vertical line indicating the rest interval. Blue lines represent the fitted exponential models. RMSE values for Block 1 and 2 are displayed adjacent to the corresponding model fit.
